# A Deep Learning Approach for Transgender and Gender Diverse Patient Identification in Electronic Health Records

**DOI:** 10.1101/2023.06.07.23290988

**Authors:** Yining Hua, Liqin Wang, Vi Nguyen, Meghan Rieu-Werden, Alex McDowell, David W. Bates, Dinah Foer, Li Zhou

## Abstract

**Background:** Although accurate identification of gender identity in the electronic health record (EHR) is crucial for providing equitable health care, particularly for transgender and gender diverse (TGD) populations, it remains a challenging task due to incomplete gender information in structured EHR fields.

**Objective:** To develop a deep learning classifier to accurately identify patient gender identity using patient-level EHR data, including free-text notes.

**Methods:** This study included adult patients in a large healthcare system in Boston, MA, between 4/1/2017 to 4/1/2022. To identify relevant information from massive clinical notes and to denoise, we compiled a list of gender-related keywords through expert curation, literature review, and expansion via a fine-tuned BioWordVec model. This keyword list was used to pre-screen potential TGD individuals and create two datasets for model training, testing, and validation. Dataset I was a balanced dataset that contained clinician-confirmed TGD patients and cases without keywords. Dataset II contained cases with keywords. The performance of the deep learning model was compared to traditional machine learning and rule-based algorithms.

**Results:** The final keyword list consists of 109 keywords, of which 58 (53.2%) were expanded by the BioWordVec model. Dataset I contained 3,150 patients (50% TGD) while Dataset II contained 200 patients (90% TGD). On Dataset I the deep learning model achieved a F1 score of 0.917, sensitivity of 0.854, and a precision of 0.980; and on Dataset II a F1 score of 0.969, sensitivity of 0.967, and precision of 0.972. The deep learning model significantly outperformed rule-based algorithms.

**Conclusion:** This is the first study to show that deep learning algorithms can accurately identify gender identity using EHR data. Future work should leverage and evaluate additional diverse data sources to generate more generalizable algorithms.

**Graphical abstract:** 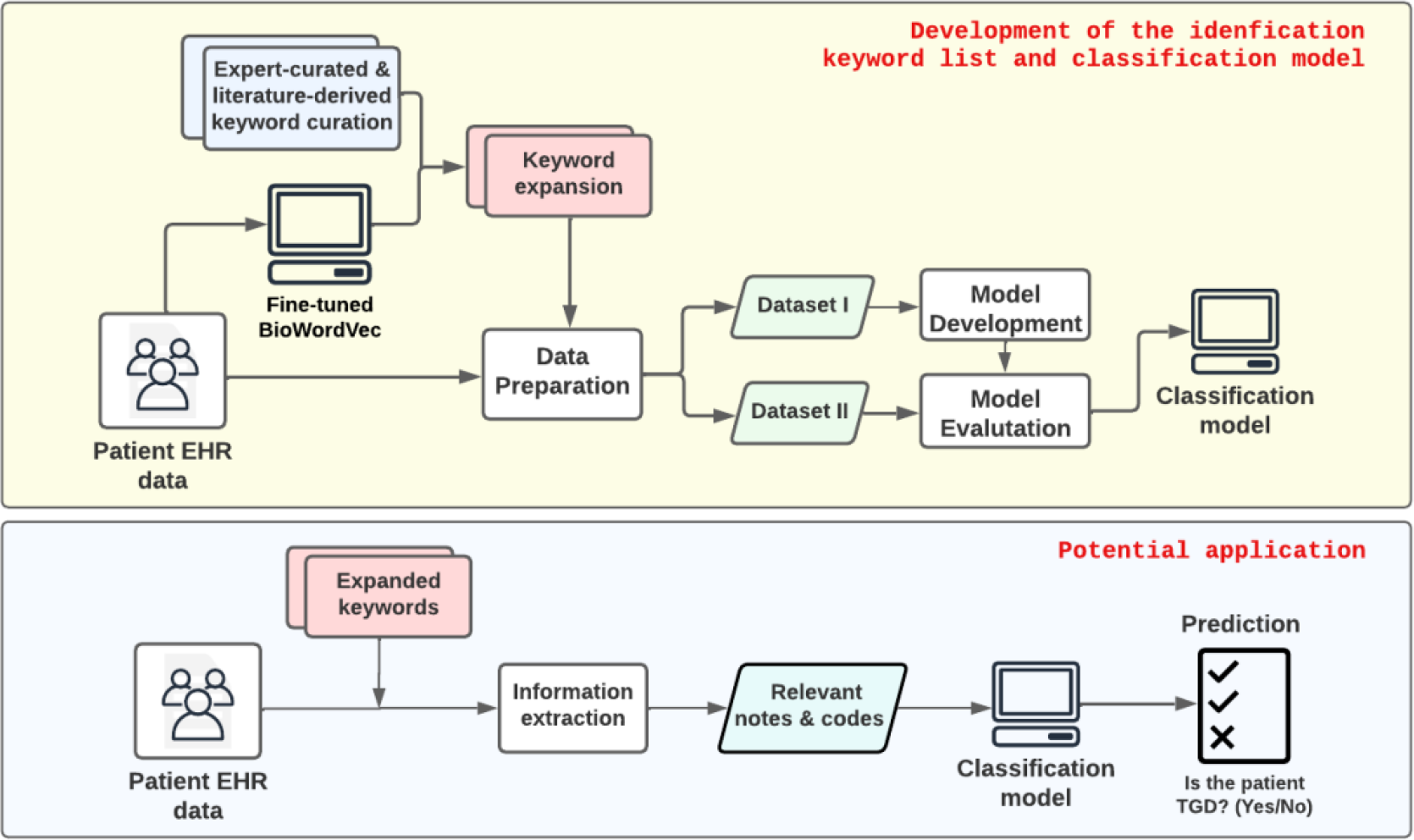

## 1. INTRODUCTION

The transgender and gender-diverse (TGD) population is growing, with estimates ranging from 0.5-4.5% among adults and 2.5-8.4% among children and adolescents [1,2]. TGD populations experience health inequities and barriers to care, and are underrepresented in research studies [3–5].

Accurate and complete sex and gender data in electronic health records (EHR) is broadly recognized as a prerequisite for improving patient safety and advancing health equity for TGD populations [6]. However, structured sex and gender information are commonly missing in EHR data, which impedes patient safety efforts and prevents high-quality TGD health research using EHR data [7–9]. Despite missingness in structured fields, detailed information about a patient’s gender identity may be available in free-text notes. Therefore, there is an urgent need to develop effective and efficient methods to identify TGD individuals within the EHR system.

Prior studies on methods to identify TGD individuals in EHR clinician notes have relied on rule-based natural language processing (NLP) algorithms that utilize a narrow set of medical codes and gender-related keywords [10–15]. Although rule-based methods are generally easier to understand and implement quickly, they may have lower accuracy than more sophisticated approaches using technology like artificial intelligence due to the difficulty of identifying complex patterns in human languages [5,8]. Pure keyword-based searches may also miss important contextual information in clinical notes, leading to false negatives.

Deep learning techniques, which are among the most sophisticated types of artificial intelligence and involve the utilization of neural networks for the analysis of large datasets, have demonstrated significant potential in clinical information studies [16,17]. These techniques are capable of learning intricate patterns and relationships within data, surpassing traditional machine learning methods in various tasks [18–20]. Furthermore, deep learning-powered NLP uses text representations to harness the wealth of information in clinical notes, making it a popular choice in patient cohort identification in EHR systems [21,22]. However, deep learning models must often overcome limitations related to data noise (such as irrelevant or inconsistent data across a patient’s EHR) and extensive annotation requirements [23–26]. Manual annotation to support sentence level prediction [27] addresses some of these challenges, but is inefficient, costly, and lacks scalability.

The objective of this study was to develop a robust deep learning-aided pipeline that leverages both structured EHR data and free-text notes for identifying TGD individuals. Through this automated approach, we aimed to reduce resource utilization, improve efficiency, and increase accuracy; the resulting applications may improve researchers’ ability to identify samples of TGD individuals in EHR data.

## 2. MATERIALS AND METHODS

### 2.1. Clinical Setting and Data Sources

This study was conducted at Mass General Brigham (MGB), a large healthcare delivery system in the Northeastern United States. The study population included patients aged ≥18 years with at least one encounter at the health system between April 1, 2017, and April 1, 2022. Patient EHR data were retrieved from MGB’s two clinical databases: the Research Patient Data Registry (RPDR) and the Enterprise Data Warehouse (EDW), which together encompass patient demographics, healthcare encounters, problem lists, billing and encounter diagnoses, procedures, and clinical notes. **A.1** details terminology used in this study. Patients who identified as ";chose not to disclose"; for gender identity or sex assigned at birth in the structured sex and gender demographic fields were excluded from the study for ethical considerations (**A.2)**.

### 2.2. Overview of the Workflow

**Figure 1** illustrates the workflow for developing and evaluating a deep learning-aided pipeline for TGD patient identification, which consisted of three steps. First, we compiled a comprehensive list of TGD keywords from three sources (expert input, published literature, and a BioWordVec model) to extract relevant information from the EHR. Next, using the keywords, we created a “screening cohort” from MGB’s EHR and split this cohort into potential non-TGD and TGD patients for model development and evaluation. For model development, we further created a balanced training dataset by leveraging an internally generated TGD patient cohort previously confirmed by clinicians. Finally, we trained a deep learning-based TGD classifier compared with several machine learning algorithms. We evaluated the effectiveness of each component of our pipeline.

**Figure 1.**
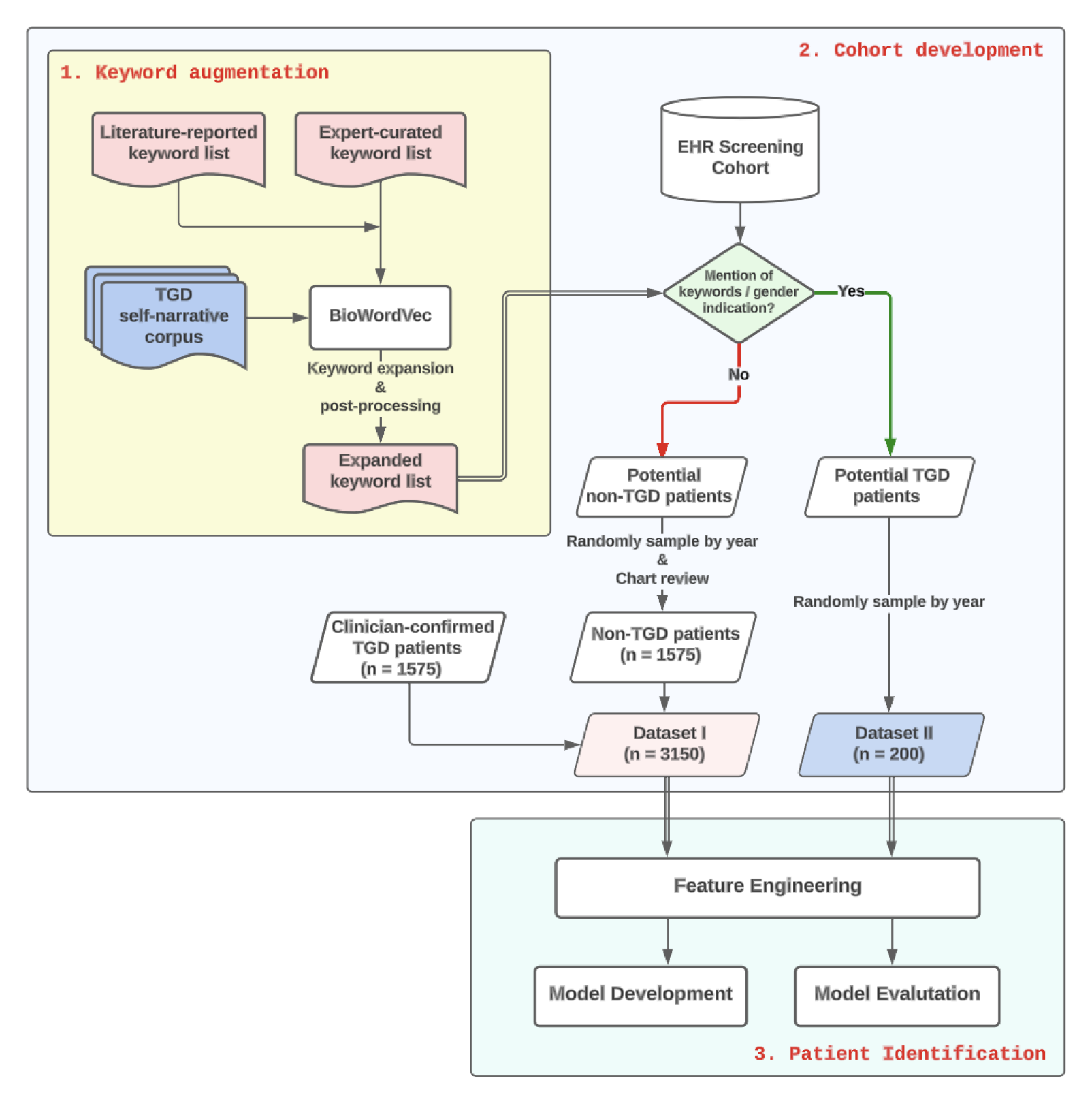
*Transgender and gender diverse identification algorithm pipeline*

### 2.3. TGD Keyword Identification

Developing keyword lists is a crucial step when building input corpora for deep learning models from patient-level EHR data. As the length of the input increases, so does the computation time and data noise. In our case, we selected the BERT architecture, which presents a maximum sequence length limitation of 512 tokens. To ensure that the most significant information is retained within this limit and with minimal noise, a list of keywords was compiled to pre-screen patient data.

To meet our objective of minimizing false negatives and maximizing prediction model accuracy, developed a comprehensive keyword list to pre-screen potential TGD individuals using three sources in sequence: expert input, published literature, and a BioWordVec model finetuned on a self-narrative corpus. Initially, a group of clinicians experienced in transgender healthcare created a list of keywords. We then identified additional keywords from relevant articles on this subject [10–12,14]. The expert-curated list and literature-reported list were then merged to form a base list, which was subsequently edited to eliminate duplicates, acronyms, and words that may introduce false positives, such as *MTF* (which is frequently used to refer to military treatment facility), *identifies as* (often followed by religious beliefs), *body dysmorphia* and *bisexual* (which are not closely related to TGD and may introduce bias), etc. In some cases, related keywords were grouped together (e.g., *transvestic disorder*, *transvestic fetish*, and *transvestite* were grouped under *transvest*), while others were not if they were not closely related to TGD or had high rates of false positives (e.g., *gender identity disorder* and *gender identity issue* were not combined with *gender identity*). Some of the keywords reflect stigmatizing terminology that was previously used to describe identities and behaviors in the TGD population, including ICD codes that have since been replaced with updated terms.

We then employed word embedding techniques to expand the base list. We used BioWordVec [28], a pre-trained word embedding model designed specifically for biomedical NLP tasks. This model used neural networks to analyze word associations in the training data and assigned each word a vector representation. For the TGD identification task, we fine-tuned the BioWordVec on a corpus of transgender-related texts [29] to create a new word embedding model. The transgender corpus contained self-narratives collected from the *asktransgender* subreddit channel. To the best of our knowledge, it is the most extensive public corpus on transgender-related topics. We then removed stop words (defined as words that carry little or no information in a language), generated unigrams, bigrams, and trigrams from the remaining text, added a new vocabulary to the BioWordVec model’s dictionary, and trained the model with three epochs.

Using the fine-tuned BioWordVec model, we extended the base list by identifying the top 30 similar phrases for each keyword in the list. Each of these phrases was manually reviewed, and those were removed if they were stop words (e.g., *hello, sis*), directly unrelated to TGD (e.g., *depression*, *anxiety*), or likely to produce false positives in keyword matching. Since the BioWordVec model was fine-tuned on a social media corpus, we further filtered it by matching the keywords against the set of clinician-verified clinical notes from TGD patients to ensure that the expanded list of keywords was relevant to our clinical context. Any keywords not appearing in notes were removed from the list.

To make the keyword list usable without deep learning models, we divided it into a main list and a complementary list. The main list’s keywords are directly TGD-related, while the complementary list contains phrases that frequently appear with TGD terms in our dataset but are less directly related to TGD, such as procedures that non-TGD patients can receive (e.g., breast augmentation, voice modification, etc.). While we separated them for the readers, we used both lists in our pipeline because it includes a BERT model, which makes predictions based on contextual information. We recommend not using the complementary list without a contextual model to avoid algorithm bias.

### 2.4. Data Preparation

#### 2.4.1. Creation of development and validation datasets

The study population comprised three groups: the cohort of clinician-confirmed TGD patients seen at the health system, potential TGD patients, and potential non-TGD patients. The latter two were selected based on the presence of TGD-related keywords in diagnoses, procedures, and notes, as well as any indication of diverse gender identity in the gender identity fields. The clinician-confirmed TGD group and the potential non-TGD group were used to create a balanced dataset for model development and evaluation, as described below. The potential TGD group was used for further evaluation of the model.

To develop and evaluate the TGD classifier, we created two datasets: Dataset I for model development, and Dataset II for further testing the model’s performance on keyword-preselected patients.

Dataset I consisted of the clinician-confirmed TGD patients as positive cases, as well as an equal number of potential non-TGD patients as negative cases. Those negative cases were randomly sampled by year among all potential non-TGD patients. We conducted a manual chart review, detailed below, on 150 randomly selected non-TGD cases and found that 146 (97.3%) were confirmed to be non-TGD patients; the remaining four patients did not have sufficient records for assessment.

Dataset II consists of a randomly selected sample of 200 potential TGD patients and was used to evaluate the ability of the trained model to predict gender identity on the remainder of the dataset.

#### 2.4.2. Chart review

A manual chart review was conducted to provide gold-standard labels of gender identity (TGD or non-TGD). The review was performed by two authors (Y.H. and V.N.) and consisted of examining demographic fields, progress notes, diagnoses and procedures, and problem lists related to gender within the EHR to determine TGD labels. Any discrepancies between the two reviewers were adjudicated by a third reviewer (D.F.). The purpose of the manual chart review was to provide accurate labels for use in training and evaluating the TGD classifier.

#### 2.4.3. Generating corpora

We extracted structured and unstructured EHR data for individual patients and converted it into a free-text format suitable for use with deep learning algorithms. However, the BERT model has a limited processing capacity, typically processing up to 512 tokens. As patient notes often exceed this limit, we employed several strategies commonly seen in various text pre-processing tasks [27,30] to shorten note length. In detail, we extracted sentences containing at least one keyword, removed duplicate sentences, and concatenated the remaining sentences in their original order. Then, regular expressions were used to remove unrelated information such as dates, times, patient identifiers, zip codes, numbers with more than three digits, parentheses and their contents, and known health system locations, such as hospital names and locations. If the notes were still longer than 400 words, we segmented the text into sentences and selected as many sentences as possible in their original order within 400 words. This process allowed us to classify the gender identity of patients based on their EHR data using the BERT model.

To incorporate structured EHR data (such as the sex and gender demographic fields, diagnoses, and procedures) into the deep learning pipeline for TGD identification, we converted the structured data into free text. This was done by inserting the names and values of the data into template sentences and then concatenating the resulting text with the processed notes. For example, the template sentences for diagnoses and procedures could be in the following format:

1. The patient was diagnosed with: DIAGNOSIS1, DIAGNOSIS2, …, DIAGNOSISn.
2. The patient received: PROCEDURE1, PROCEDURE2, …, PROCEDUREm.

Here, DIAGNOSIS and PROCEDURE are unique names of the diagnosis and procedure code, and n and m are the number of diagnoses and procedures, respectively.

Similarly, for we converted patient sex and gender demographic field information using the template sentence: “The patient’s sex at birth is SEX_AT_BIRTH; legal sex is LEGAL_SEX; gender identity is GENDER_IDENTITY”.

Our final corpus for the model consists of patient notes concatenated from sentences of individual patients in the following order: 1) diagnoses and procedures, 2) sex and gender demographics, and 3) extracted note sentences.

### 2.5. Classification Models

#### 2.5.1. Deep learning-based classifier

To classify patients as transgender or cisgender, we built a linear classification model, in which we used Bio_ClinicalBERT [31], a variant of bidirectional encoder representations from transformers (BERT) [32] that has been further trained on extensive biomedical data, to encode the processed patient notes. We added a linear classifier after the ClinicalBERT embedding layers and froze all but the last layer to prevent overfitting. Then, we trained the model on a binary classification task of identifying transgender patients, utilizing binary cross-entropy loss.

Given the small size of our data sets, we froze all but the last three layers of the Bio_ClinicalBERT model before fine-tuning. We set the maximum length of tokens to 512, and both the training and validation batch sizes to 8. The model was trained using a learning rate of 3e-5 for four epochs, and its performance was evaluated every 100 training steps. Training and validation typically took 8 to 10 minutes on an NVIDIA Quadro p6000 GPU with 24 GB of memory.

#### 2.5.2. Baselines

We conducted several experiments to evaluate the performance of our model by comparing it with several baseline approaches, including rule-based and statistical machine learning algorithms. While a few studies have reported the use of rule-based approaches in TGD identification, none of them, to the best of our knowledge, have used machine learning-based approaches. For the rule-based approaches, we used the best single-and combined-rule algorithm proposed by Guo et al [14].

We also tested traditional statistical machine learning methods. We transformed the text into n-grams (unigrams, bigrams, and trigrams) and used the term frequency-inverse document frequency (TF-IDF) [33,34], a widely adopted method to measure the relevance of n-grams to a document across a collection of documents, to encode the texts. We applied XGBoost, support vector machine (SVM), random forest, and logistic regression to classify the encoded texts using the Scikit-learn package. The parameters of the classic machine learning models were optimized using a grid search on the training set.

Additionally, to evaluate the performance of the keyword expansion module, we compared the pipeline’s performance in two different settings: (1) using only the baseline keyword list (i.e., literature-reported keywords and expert-curated keywords), and 2) using the expanded keyword list, but without the classification module. We considered patients with any keyword matches in any data fields (e.g., notes, diagnosis, and procedures) as TGD.

#### 2.5.4. Evaluation metrics and strategy

We used the F1 score, a metric that combines precision and recall, to evaluate the performance of our model. In addition to the F1 score, we also report the mean and standard deviation of sensitivity, specificity, positive predictive value (PPV), negative predictive value (NPV), accuracy, area under the receiver operating characteristic curve (AUROC), and area under the precision-recall curve (AUPRC), which were calculated based on five-fold cross-validation. This allowed us to assess the stability and robustness of our results and to determine the overall performance of the model.

To ensure the model could accurately predict the gender identity of patients with missing structured gender demographic information, we conducted a sub-analysis to assess the model’s performance on a subset of patients who had missing values in the gender fields in both the development and evaluation datasets. Only patients whose gender identity values were “unknown” were included in this sub-analysis. Patients with a “chose to not disclose” value were excluded, consistent with the main analysis.

We also conducted an error analysis on both datasets to gain a deeper understanding of where the model is likely to fail. Specifically, we aimed to identify the types of errors made by the model as well as the characteristics of the patients for whom the model demonstrated poor performance.

## 3. RESULTS

### 3.1. TGD Keyword Identification

The expert-curated keyword list contained 27 keywords and the literature-reported keyword list contained 53 keywords (**table 1**). After merging the two lists and removing any misused keywords, there were 51 unique keywords. Following keyword expansion, the total number of keywords in the expanded list reached 364. Among these, 109 (29.9%) keywords—including 58 novel ones—were referenced at least once in the clinical notes of the study group, and thus were incorporated into our final expanded keyword list.

**Table 1.**
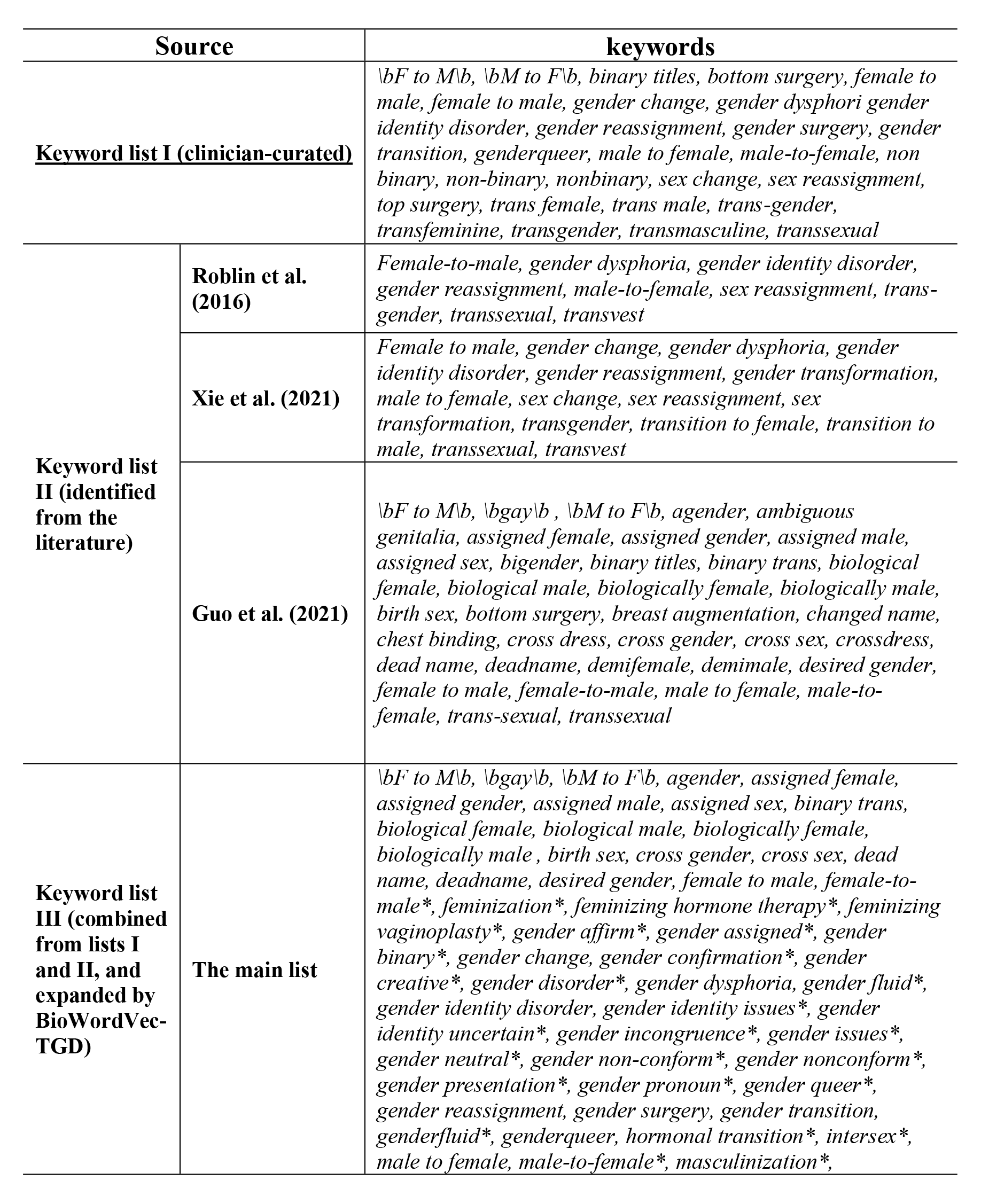
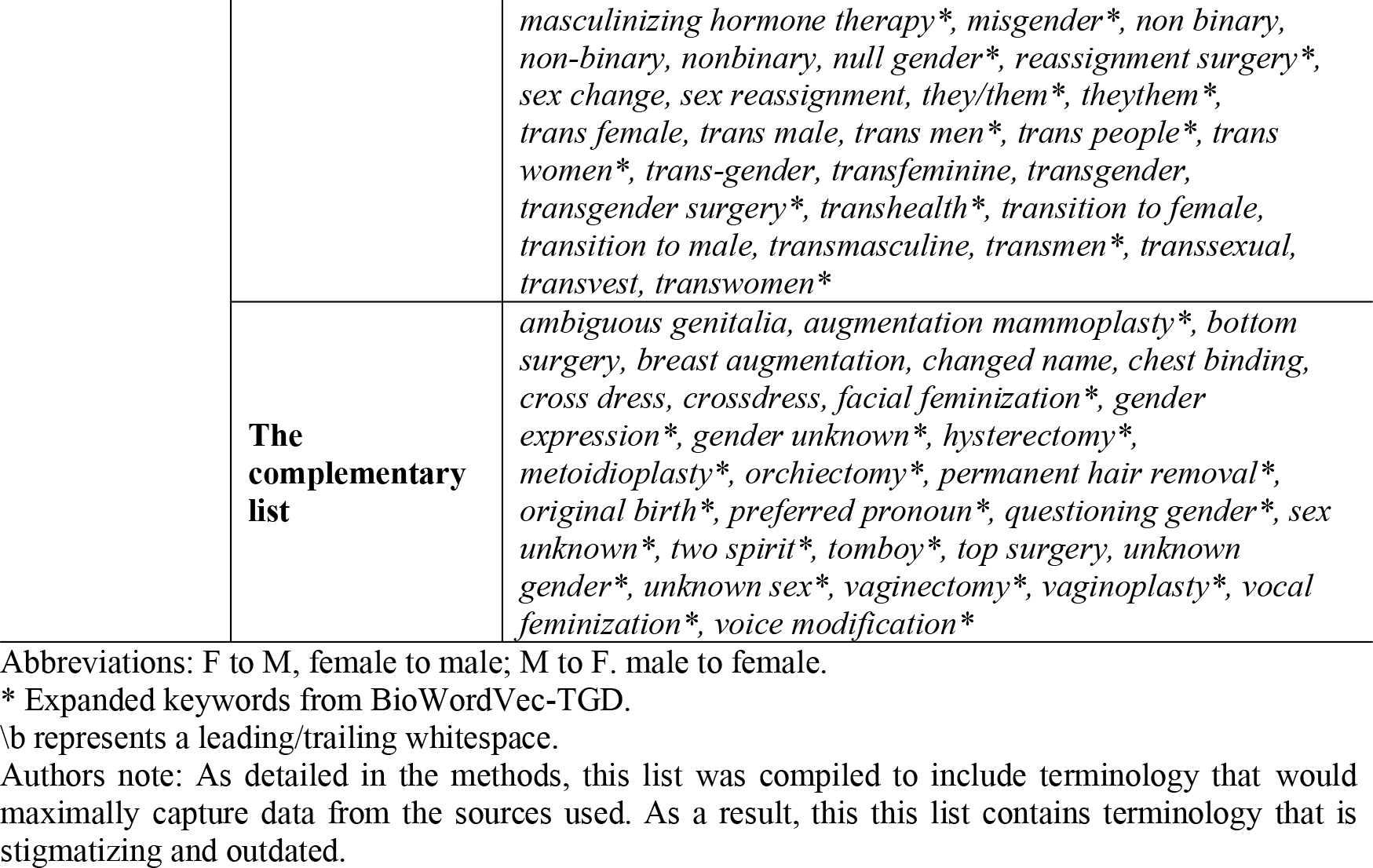
*TGD-related Keyword lists*

| Source |  | keywords |
| --- | --- | --- |
| **<u>Keyword list I (clinician-curated)</u>** |  | *\bF to M\b, \bM to F\b, binary titles, bottom surgery, female to male, female to male, gender change, gender dysphoria, gender identity disorder, gender reassignment, gender surgery, gender transition, genderqueer, male to female, male-to-female, non binary, non-binary, nonbinary, sex change, sex reassignment, top surgery, trans female, trans male, trans-gender, transfeminine, transgender, transmasculine, transsexual* |
| **Keyword list II (identified from the literature)** | **Roblin et al. (2016)** | *Female-to-male, gender dysphoria, gender identity disorder, gender reassignment, male-to-female, sex reassignment, trans-gender, transsexual, transvest* |
|  | **Xie et al. (2021)** | *Female to male, gender change, gender dysphoria, gender identity disorder, gender reassignment, gender transformation, male to female, sex change, sex reassignment, sex transformation, transgender, transition to female, transition to male, transsexual, transvest* |
|  | **Guo et al. (2021)** | *\bF to M\b, \bgay\b, \bM to F\b, agender, ambiguous genitalia, assigned female, assigned gender, assigned male, assigned sex, bigender, binary titles, binary trans, biological female, biological male, biologically female, biologically male, birth sex, bottom surgery, breast augmentation, changed name, chest binding, cross dress, cross gender, cross sex, crossdress, dead name, deadname, demifemale, demimale, desired gender, female to male, female-to-male, male to female, male-to-female, trans-sexual, transsexual* |
| **Keyword list III (combined from lists I and II, and expanded by BioWordVec-TGD)** | **The main list** | *\bF to M\b, \bgay\b, \bM to F\b, agender, assigned female, assigned gender, assigned male, assigned sex, binary trans, biological female, biological male, biologically female, biologically male, birth sex, cross gender, cross sex, dead name, deadname, desired gender, female to male, female-to-male\*, feminization\*, feminizing hormone therapy\*, feminizing vaginoplasty\*, gender affirm\*, gender assigned\*, gender binary\*, gender change, gender confirmation\*, gender creative\*, gender disorder\*, gender dysphoria, gender fluid\*, gender identity disorder, gender identity issues\*, gender identity uncertain\*, gender incongruence\*, gender issues\*, gender neutral\*, gender non-conform\*, gender nonconform\*, gender presentation\*, gender pronoun\*, gender queer\*, gender reassignment, gender surgery, gender transition, genderfluid\*, genderqueer, hormonal transition\*, intersex\*, male to female, male-to-female\*, masculinization\*,* |

|  |  |
| --- | --- |
|  | *masculinizing hormone therapy\**, *misgender\**, *non binary*, *non-binary*, *nonbinary*, *null gender\**, *reassignment surgery\**, *sex change*, *sex reassignment*, *they/them\**, *theythem\**, *trans female*, *trans male*, *trans men\**, *trans people\**, *trans women\**, *trans-gender*, *transfeminine*, *transgender*, *transgender surgery\**, *transhealth\**, *transition to female*, *transition to male*, *transmasculine*, *transmen\**, *transsexual*, *transvest*, *transwomen\** |
| **The complementary list** | *ambiguous genitalia*, *augmentation mammoplasty\**, *bottom surgery*, *breast augmentation*, *changed name*, *chest binding*, *cross dress*, *crossdress*, *facial feminization\**, *gender expression\**, *gender unknown\**, *hysterectomy\**, *metoidioplasty\**, *orchiectomy\**, *permanent hair removal\**, *original birth\**, *preferred pronoun\**, *questioning gender\**, *sex unknown\**, *two spirit\**, *tomboy\**, *top surgery*, *unknown gender\**, *unknown sex\**, *vaginectomy\**, *vaginoplasty\**, *vocal feminization\**, *voice modification\** |

Abbreviations: F to M, female to male; M to F, male to female.
\* Expanded keywords from BioWordVec-TGD.
\b represents a leading/trailing whitespace.
Authors note: As detailed in the methods, this list was compiled to include terminology that would maximally capture data from the sources used. As a result, this this list contains terminology that is stigmatizing and outdated.

### 3.2. Dataset Characteristics

Dataset I contained 3,150 patients, of whom 1575 (50%) were clinician-confirmed TGD patients. Dataset II contained 200 patients, of which 180 (90%) were TGD patients. **Table 2** displays the key characteristics of the datasets as well as the TGD and non-TGD patients in each dataset. TGD keywords were more frequently identified in clinical notes than in the diagnosis field, while the procedure field had the lowest frequency. For example, TGD keywords were mentioned in 89.02% of the TGD patients’ notes in Dataset I and 95.56% of the TGD patients’ notes in Dataset II. In contrast, in Dataset I, keywords were only mentioned in 60.76% and 26.8% of the TGD patients’ diagnoses and procedures, respectively. Similarly, in Dataset II, only 103 (57.22%) TGD patients had keywords in their diagnosis fields, and 10 (5.56%) in procedure fields. Out of 200 randomly selected patients with keyword matches, 20 were found to be non-TGD. Among these false positives, 3 (15%) had procedure matches, and 17 (85%) had note matches. We identified high missingness in the structured gender demographic fields: in Dataset I, 1247 (39.59%) patients had missing gender identity values, and in Dataset II, 99 (49.5%) patients had missing values.

**Table 2.** *Summary of two datasets for model development and evaluation*

|  | **Dataset I**<br>(N = 3150) |  | **Dataset II**<br>(N = 200) |  |
| --- | --- | --- | --- | --- |
|  | **Clinician-confirmed TGD patients**<br>(N=1575)<br>n (%) | **Non-TGD patients filtered by keyword search**<br>(N=1575)<br>n (%) | **TGD patients by chart review**<br>(N=180)<br>n (%) | **Non-TGD patients by chart review**<br>(N=20)<br>n (%) |
| **Age, mean (SD) year** | 35.94 (16.04) | 60.92 (18.0) | 34.52 (15.48) | 57.85 (20.27) |
| **Race, n (%)** |  |  |  |  |
| Asian | 77 (4.89) | 37 (2.35) | 8 (4.44) | 1 (5.0) |
| Black | 116 (7.37) | 84 (5.33) | 12 (6.67) | 2 (10.0) |
| More than one race | 50 (2.54) | 6 (0.38) | 6 (3.33) | 0 (0.0) |
| Other | 177 (11.24) | 116 (7.37) | 24 (13.33) | 2 (10.0) |
| White | 1155 (73.33) | 1332 (84.57) | 130 (72.22) | 15 (75.0) |
| **Ethnicity** |  |  |  |  |
| Hispanic | 22 (1.40) | 41 (2.60) | 9 (5.0) | 1 (5.0) |
| Non-Hispanic | 1351 (85.78) | 1321 (83.87) | 146 (81.11) | 15 (75.0) |
| Other | 415 (12.83) | 213 (13.52) | 25 (13.89) | 4 (20.0) |
| **Patients with keywords, n (%)** |  |  |  |  |
| Diagnoses | 957 (60.76) | 0 | 103 (57.22) | 0 |
| Procedures | 422 (26.8) | 0 | 10 (5.56) | 3 (15.0) |
| Clinical notes | 1402 (89.02) | 0 | 172 (95.56) | 17 (85.0) |
| **Patients with missing gender fields, n (%)** | 884 (56.13) | 691 (43.87%) | 84 (46.67) | 15 (75.0) |

### 3.3. Model Performances on Dataset I

**Table 3** shows the performance of our models on dataset I. *Bio_ClinicalBERT_TGD* achieved an F1 score of 0.917, a sensitivity of 0.854, and a precision of 0.980, which significantly outperformed the rule-based baseline algorithms. Compared to other machine learning algorithms, *Bio_ClinicalBERT_TGD* achieved slightly better performance, with an AUROC of 0.913 (95% CI, 0.891, 0.935) and an AUPRC of 0.956 (95% CI, 0.941, 0.970).

**Table 3.** *Performance of TGD identification algorithms on Dataset I (development set)*

|  |  | F1 | Sensitivity | Specificity | Precision | NPV | Accuracy | AUROC (95% CI) | AUPRC (95% CI) |
| --- | --- | --- | --- | --- | --- | --- | --- | --- | --- |
| **Rule-based** | Exact Match | 0.586 | 0.980 | 0.962 | 0.730 | 0.728 | 0.796 | N/A | N/A |
|  | Augmented Match | 0.857 | 0.883 | 0.882 | 0.858 | 0.869 | 0.870 | N/A | N/A |
|  | Guo et al. (single)<sup>1*</sup> | 0.816 | 0.723 | 0.952 | 0.936 | 0.777 | 0.838 | N/A | N/A |
|  | Guo et al. (combined)<sup>2*</sup> | 0.843 | 0.766 | 0.951 | 0.939 | 0.804 | 0.859 | N/A | N/A |
| **Machine Learning** | Random Forest | 0.892 | 0.832 | 0.976 | 0.972 | 0.860 | 0.904 | 0.904 (0.880, 0.926) | 0.944 (0.926, 0.959) |
|  | Support Vector Machine | 0.886 | 0.808 | 0.993 | 0.991 | 0.844 | 0.900 | 0.900<br>(0.876, 0.923) | 0.947<br>(0.932, 0.961) |
|  | Linear Regression | 0.882 | 0.799 | **0.994** | **0.991** | 0.837 | 0.896 | 0.896<br>(0.872, 0.919) | 0.946<br>(0.931, 0.959) |
|  | XGBoost | 0.892 | 0.828 | 0.978 | 0.975 | 0.858 | 0.903 | 0.903<br>(0.879, 0.926) | 0.945<br>(0.927, 0.960) |
| **Deep Learning** | Bio_ClinicalBERT_TGD | **0.917** | **0.854** | 0.983 | 0.980 | **0.865** | **0.912** | **0.913**<br>**(0.891, 0.935)** | **0.956**<br>**(0.941, 0.970)** |

<sup>1</sup>Best single-rule algorithm was based on $\geq 2$ diagnosis codes and $\geq 1$ keyword(s)
<sup>2</sup>Best combined rule was either gender field indicates transgender or $\geq 1$ diagnosis code(s) plus $\geq 1$ TGD keyword(s)
\*Codes and keywords can be found in the paper by Guo et al. [17].

The augmented match algorithm, which relies on a single rule based on the presence or absence of any keywords, achieved an F1 score of 0.857 and a sensitivity of 0.883, outperforming previously published best-combined rules approach in [14].

Finally, traditional machine learning classifiers on TF-IDF encoded text features had comparable performance to *Bio_ClinicalBERT_TGD*, with only a 0.2 to 0.3 sacrifice in F1.

**Table 4** presents the algorithms’ performance on the subset of patients from Dataset I with missing structured gender field values. *Bio_ClinicalBERT_TGD* remained the best-performing model, achieving the highest F1 score of 0.923, the highest sensitivity of 0.906 and AUROC of 0.940. *Bio_ClinicalBERT_TGD* significantly outperformed the rule-based algorithms in terms of F1 score, sensitivity, specificity, precision, NPV, and accuracy. *Bio_ClinicalBERT_TGD* slightly outperformed machine learning models in all the metrics except specificity and precision. The rule-based baseline models all showed a decrease in performance compared to the performance in the entire Dataset I. In contrast, the machine learning and deep learning models showed a slight improvement in performance.

**Table 4.** *Sub-analysis of patients with missing structured sex and gender demographics in Dataset I*

|  |  | F1 | Sensitivity | Specificity | Precision | NPV | Accuracy | AUROC<br>(95% CI) | AUPRC<br>(95% CI) |
| --- | --- | --- | --- | --- | --- | --- | --- | --- | --- |
| **Rule-based** | Exact Match | 0.254 | 0.983 | 0.852 | 0.770 | 0.391 | 0.777 | N/A | N/A |
|  | Augmented Match | 0.658 | 0.908 | 0.756 | 0.860 | 0.703 | 0.833 | N/A | N/A |
|  | Guo et al. (single) | 0.766 | 0.674 | 0.951 | 0.887 | 0.837 | 0.851 | N/A | N/A |
|  | Guo et al. (combined) | 0.788 | 0.706 | 0.951 | 0.892 | 0.850 | 0.862 | N/A | N/A |
| **Machine Learning** | Random Forest | 0.901 | 0.837 | 0.957 | 0.977 | 0.728 | 0.874 | 0.897<br>(0.870, 0.923) | 0.963<br>(0.949, 0.975) |
|  | Support Vector Machine | 0.900 | 0.827 | **0.979** | **0.988** | 0.721 | 0.874 | 0.903<br>(0.878, 0.926) | 0.967<br>(0.956, 0.977) |
|  | Linear Regression | 0.889 | 0.811 | 0.971 | 0.984 | 0.701 | 0.861 | 0.891<br>(0.865, 0.916) | 0.962<br>(0.950, 0.973) |
|  | XGBoost | 0.901 | 0.837 | 0.957 | 0.977 | 0.728 | 0.874 | 0.897<br>(0.870, 0.923) | 0.963<br>(0.949, 0.975) |
| **Deep Learning** | Bio_ClinicalBERT_TGD | **0.923** | **0.906** | 0.975 | 0.940 | **0.960** | **0.954** | **0.940**<br>**(0.912, 0.964)** | **0.937**<br>**(0.908, 0.961)** |

<sup>1</sup>Best single-rule algorithm was based on $\geq 2$ diagnosis codes and $\geq 1$ keyword(s)
<sup>2</sup>Best combined rule was either gender field indicates transgender or $\geq 1$ diagnosis code(s) plus $\geq 1$ TGD keyword(s)

### 3.4. Bio_ClinicalBERT_TGD on Dataset II

**Table 5** shows *Bio_ClinicalBERT_TGD*’s performance on Dataset II, the patients randomly sampled from the potential TGD patient group (**Figure 1**). *Bio_ClinicalBERT_TGD* had an F1 score of 0.977, with a higher sensitivity of 0.967 and a higher precision of 0.988 compared to its performance on Dataset I. The mode’s specificity and NPV dropped to 0.80 and 0.75, respectively, indicating that it was better at identifying true positive cases than true negative cases.

**Table 5.** *Performance of Bio_ClinicalBERT_TGD on Dataset II.*

|  | **F1** | **Sensitivity** | **Specificity** | **Precision** | **NPV** | **Accuracy** | **AUROC**<br>**(95% CI)** | **AUPRC**<br>**(95% CI)** |
| --- | --- | --- | --- | --- | --- | --- | --- | --- |
| All patients | 0.977 | 0.967 | 0.900 | 0.988 | 0.750 | 0.960 | 0.858<br>(0.755, 0.954) | 0.984<br>(0.970, 0.996) |
| Patients with missing structured sex and gender demographics | 0.970 | 0.976 | 0.800 | 0.964 | 0.857 | 0.939 | 0.865<br>(0.770, 0.960) | 0.988<br>(0.974, 1.000) |

In the sub-analysis set of patients missing structured gender demographics, the model experienced a 0.007 decrease in the F1 score. The NPV increased to 0.857, suggesting that among patients with missing structured gender demographic data, the model achieved better balance in predicting true positive and true negative cases.

### 3.5. Error Analysis

A manual chart review of the false classifications by Bio_ClinicalBERT_TGD on Datasets I and II was conducted to summarize the root causes behind the false positives and negatives (Figure 2).

**Figure 2.**
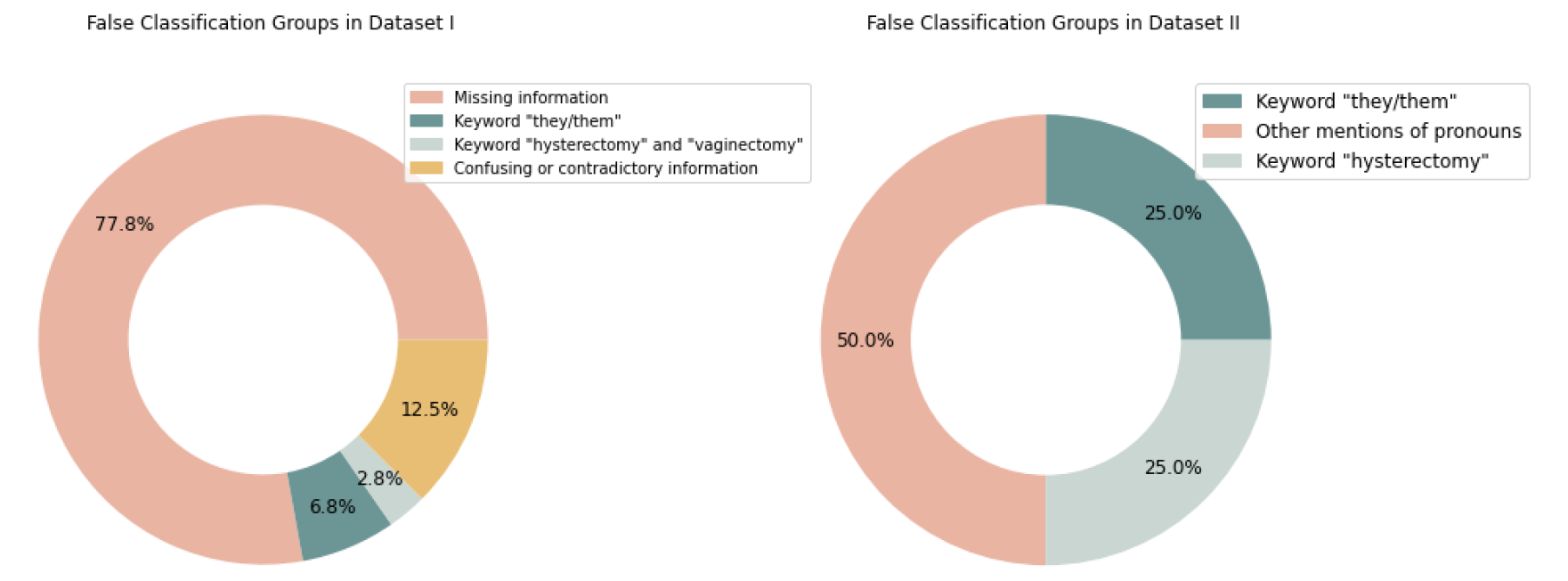
*Error analysis for false classification groups in Dataset I and II. Dataset I had 149 false negatives and 39 false positives. Dataset II had six false negatives and two false positives.*

For Dataset I, which consisted of five validations, 149 false negatives and 39 false positives were found. Most false negatives (91.95%, n=137) were attributed to an absence of sufficient information to conclusively determine a patient’s gender identity. This issue primarily arose in cases where patients had not selected any notes and the available sex and gender demographics were insufficient for accurate identification. A further 12 patients (8.05%) were identified via the pronoun ";*they/them*"; but the model failed to predict their gender, likely due to an inadequate number of training samples containing these pronouns.

The false positives in Dataset I were mainly triggered by keywords found within a complementary list. Three instances highlighted the sole keyword “hysterectomy” and two instances presented the keyword “vaginectomy.” This suggests a misinterpretation by the model, inferring a likely TGD identity for patients who had undergone a hysterectomy or vaginectomy. This bias may be the result of insufficient negative training samples containing details about these procedures, causing the model to form an overgeneralized association between these procedures and TGD identities. Additional false positives were found with confusing or contradictory information. For example, three patients had gender identity listed as unknown in structured demographics but mentioned they were biologically female or make in notes; one instance contained contradictory information, with the sex assigned at birth recorded as “male,” while the patient note indicated “biologically female.";

Dataset II yielded six false negatives and two false positives. All false negatives were related to evidence from pronouns: two instances were unable to correctly associate ";preferred pronouns: they/them"; with TGD individuals, three instances contained ";preferred pronouns: she they,"; and one instance showed “preferred pronouns are: he/him, they/their”. Both false positives were associated with mentions of hysterectomy.

## 4. DISCUSSION

In this study, we developed an accurate and efficient method for transgender and gender diverse identification in an EHR. In doing so, we were able to overcome some of the limitations of prior methods that relied on structured EHR data and rule-based algorithms. Overall, identification of this group has been a difficult problem which needs to be solved to deliver better care to these populations. We were able to develop multiple classification models, based on different machine learning-based NLP approaches, that leverage rich clinical data to achieve high performance.

This study represents a significant advancement in the identification of TGD individuals in EHRs by pioneering the use of machine learning to aid the process. The robust deep learning-aided pipeline effectively outperforms the previously predominant methodologies which relied on rule-based algorithms and a limited set of gender-related keywords and medical codes. These conventional methodologies often were limited in accuracy and lacked the pattern recognition capabilities inherent in deep learning techniques. We specifically benchmarked our models against the work of Guo et al. [14], a previously published comprehensive TGD phenotyping and identification work. Our results indicate that our algorithms consistently outperform their best rule-based approaches, thereby demonstrating the tangible benefits of our deep learning application in TGD identification.

The research pipeline we constructed, which includes a broad keyword list and multiple machine learning models, made a substantial contribution to the superb performance of gender identity detection. Across all metrics—F1 scores, accuracy, sensitivity, precision, PPV, and NPV—our methods excelled in both datasets compared to rule-based baselines. Algorithm evaluation across two datasets and two sub-analyses on patients without explicit sex and gender demographics demonstrated the superior accuracy of our machine learning-based algorithms. Moreover, they proved to be less vulnerable to gaps in sex and gender demographics, demonstrating their robustness in the face of data scarcity. Notably, the pipeline proved to be feasible and stable in classifying patient gender at the patient level, which is widely recognized as the most challenging level for prediction. Moreover, it is adaptable to note-, section-, or sentence-level predictions, although these levels require more labeling work. In doing so, our work helps to overcome a major barrier to EHR-based tools for population-level research and patient-level care, particularly given the large missing data in structured sex and gender fields. Specifically, these models could provide more complete information for downstream tasks that already rely on the gender fields, such as laboratories, rooming modules, preventative screening, population health programs, risk calculators and other applications. Future studies may compare patient-oriented outcomes in these areas using these models compared to current methods.

In addition to the Bio_ClinicalBERT_TGD model, our experiments indicated that random forest and XG-Boost, using TF-IDF encoded text features as input, also performed reasonably well on Dataset I and the sub-analysis. While BERT models are generally considered state-of-the-art for text classification tasks, they may not always be the most practical solution due to their high computational requirements and the need for large amounts of training data. In contrast, random forest and XG-Boost have lower computational resource requirements and faster computation speeds, which make them more suitable for classifying large numbers of patients in the EHR database. Depending on the specific needs and available resources, these traditional machine learning models could be a suitable alternative to BERT.

Our literature review of TGD identification enabled us to detect and correct several inaccuracies in previous conventions. We removed terms and acronyms that could result in erroneous diagnoses from the literature-reported list, such as ";MTF (male to female)";, ";identifies as";, ";body dysmorphia";, and ";bisexual";. In our examination, ";MTF"; is often used to denote Military Treatment Facilities in clinical notes, while ";identifies as"; is commonly linked to religious convictions, and the last two terms are not strongly associated with TGD. Additionally, we observed that acronyms are typically employed after the full term has been introduced. Finally, we partitioned our keyword list into a primary and supplementary list, acknowledging that the supplementary list may lead to a high rate of false positives and emphasizing the importance of sufficient training data to differentiate between complementary keywords and definite indications of TGD. Together, these efforts support portability and generalizability.

The generalizability of deep learning models is largely limited due to Health Insurance Portability and Accountability Act (HIPAA) restrictions on sharing labeled patient-level data. To overcome this limitation, our method incorporates a partially reusable component, specifically the keyword extension for data denoising, which can be applied across different institutions. Furthermore, the model-building process in our approach is designed to be straightforward, allowing for easy implementation and adaptation in various healthcare settings. Lastly, our approach can be applied to other case identification and phenotyping tasks using her data.

## 5. LIMITATIONS

Our study has several limitations that need to be acknowledged. Firstly, the BioWordVec model used to generate TGD keywords was primarily trained on PubMed data and social media posts. As a result, it might be biased towards these data sources and may not capture a complete set of keywords used in clinical notes. This limitation could have affected the model’s ability to accurately identify and classify TGD-related content in clinical notes. Secondly, our study relied on training and test sets from a single institution, which lacks external validity. Future research could benefit from utilizing larger and more diverse datasets collected from multiple institutions to improve the model’s performance and validate it across different healthcare settings. Thirdly, our positive and negative samples were heterogeneous, potentially limiting the diversity of the final training set. This lack of diversity may have hindered the model’s ability to fully understand all the keywords and concepts related to TGD. Our error analysis revealed that Bio_ClinicalBERT_TGD was often confused with *hysterectomy* and *they/them*. This confusion may be attributed to the lack of training samples with the *they/them* keyword for the model to effectively learn the relationship between these pronouns and TGD, and that we excluded any keyword matches in the negative cohort to reduce labeling work. Finally, some patients did not have information in their notes that matched TGD-related information. We attempted to identify potentially relevant information using the trained *Bio_ClinicalBERT_TGD* model and a simple clustering pipeline in a previous framework [35]. However, it did not improve classification performance; more specifically designed techniques such as iteratively the most informative instances through semi-supervised learning should be investigated in future work.

## 6. CONCLUSION

We utilized machine learning-based NLP techniques that include both clinical notes and structured EHR data to identify gender identity. Better approaches to doing this will be helpful in addressing the needs of gender diverse populations. Future work should focus on addressing improving performance by incorporating additional diverse and representative data sources, increasing training and test set sizes, and ensuring balanced sample distribution models that are generalizable and actionable for the clinical domain.

### Abbreviations

BERT: Bidirectional Encoder Representations from Transformers
EHR: Electronic Health Records
MGB: Mass General Brigham
NLP: Natural Language Processing
TGD: Transgender and Gender Diverse
SVM: Support Vector Machine
TF-IDF: Term Frequency-Inverse Document Frequency

## Data Availability

The data sets used for training and evaluation in this study are available upon reasonable request from the corresponding author, pending the necessary institutional reviews and approvals.

## ACKNOWLEDGEMENTS

Credit authorship contribution statement:

**Yining Hua**: Conceptualization, Data Curation, Methodology, Implementation, Formal analysis. Writing - Original Draft & Editing. **Liqin Wang**: Methodology, Writing - Original Draft, Supervision. **Vi Nguyen**: Data Curation, Writing - Review & Editing. **Meghan Rieu-Werden**: Data curation, Writing - Review & Editing. **Alex McDowell**: Data curation, Writing-Review & Editing. **David W. Bates**: Writing - Review & Editing, Supervision. **Dinah Foer**: Conceptualization, Writing - Review & Editing, Supervision, Project administration. Funding acquisition. **Li Zhou**: Resources, Writing - Review & Editing, Supervision.

## A. Appendices

**A1.**
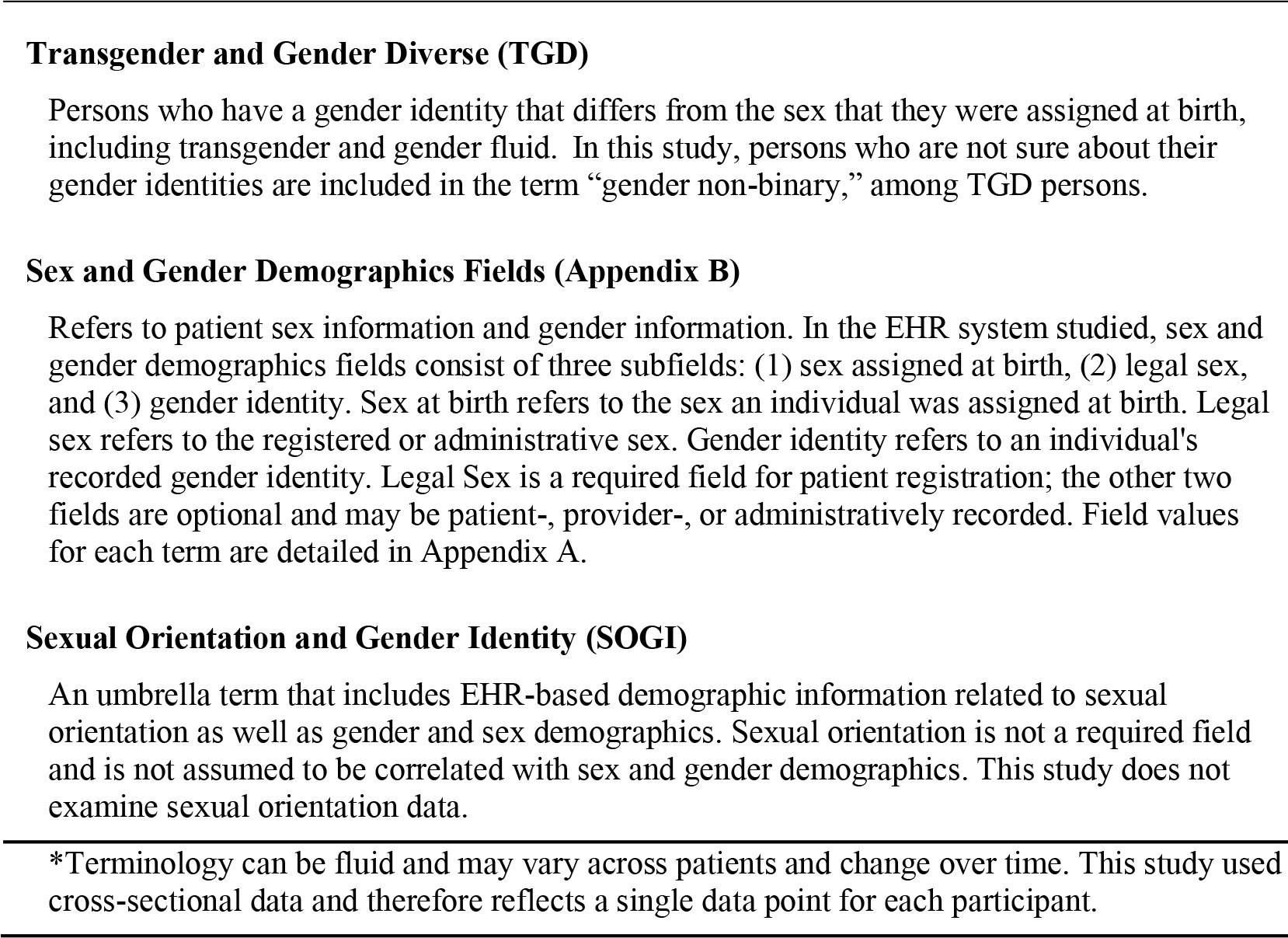
Study terminology*

**A.2.** Sex and gender demographics fields in the EHR system

A.2. Sex and gender demographics fields in the EHR system
|  |  |
| --- | --- |
| Gender (Legal Sex) | Female |
|  | Male |
|  | Unknown |
|  | X (Non-Binary)* |
| Gender Identity | Chose not to disclose |
|  | Female |
|  | Male |
|  | Non-binary* |
|  | Other* |
|  | Queer/Genderqueer* |
|  | Questioning/Unsure* |
|  | Transgender Female (Male-to-Female)* |
|  | Transgender Male (Female-to-Male)* |
|  | Unknown |
| Sex at Birth | Chose not to disclose |
|  | Female |
|  | Male |
|  | Uncertain* |
|  | Unknown |

\*Structured values considered as TGD, if available. Patient with a field value of “chose not to disclose” for gender identity or sex at birth were excluded from the study. “Unknown” was not considered a TGD indication because it does not contain determinant information. Terminology reflects the field options in the EHR at the time of data entry.

## Notes

### Competing Interest Statement

The authors have declared no competing interest.

### Funding Statement

This work was supported by a research grant from CRICO, the medical malpractice insurance organization.

### Author Declarations

Mass General Brigham IRB #2021P001964

